# Deeply-Dyed Nanobead System Based Lateral Flow Assay for Rapid Point-of-Care Drug Testing

**DOI:** 10.1101/2021.12.06.21267347

**Authors:** Lingzhi Fan, Jianbing Wu, Jing Yang, Fugang Li, Wannian Yan, Fei Tan, Madeline Zhang, Mohamed S. Draz, Huanxing Han, Pengfei Zhang

## Abstract

Point-of-care test (POCT), which allows for rapid and sensitive screening of drugs abuse, is essential and can significantly reduce the clinical, economic and social impact of the opioid crisis worldwide. However, the traditional gold nanoparticle-based lateral flow immunoassay strip is not sensitive enough for detection of trace drugs in hair sample. Herein, we aimed to develop a more specific system using a composite polymer-based nanobead that is deeply dyed with phthalocyanine or similar oil soluble dyes, and termed as deeply dyed nanobead (DDNB). The prepared composite nanobeads displayed a clear core-shell structure and the core/shell ratios were readily controlled by polymer/dye feeding ratios. The absorbance stable nanobeads capped with carboxyl groups were covalently conjugated with antibodies, and were employed for preparation of lateral flow immunoassay strips for sensitive detection of drugs in hair with naked eye. The developed platform allows the detection of drugs such as morphine and methamphetamine in hair samples within 13 min (including hair sample processing ∼5 min). The cut-off value of DDNB strip for methamphetamine detection with naked eye is down to concentration of 8.0 ng/mL, which is about 3.1 times more sensitive than the traditional gold nanoparticles based lateral flow immunoassay. Moreover, the colorful DDNB system has the potential for multiplexing detection of analytes at point-of-care settings and with low cost.

## Introduction

Drugs of abuse are a primary social and public health concern, causing severe illness and injuries to millions of people worldwide^1-3^. In 2019, over two million illicit drug abusers were documented in China^4^. To meet the requirement of rapid, sensitive, low cost, and non/minimally-invasive detection of abused drugs is of great importance for forensic testing^5-7^ and in clinical setting^8-10^ to evaluate the efficacy of medications.

Compared with drug testing in urine^11, 12^, hair testing for drug-of-abuse has become increasingly more important^5, 13, 14^. Firstly, hair testing allows for measuring a long-term (up to 90 days) drug addiction, while urine testing is only suitable for detecting recent (within 48 hours) drug use. Moreover, for on-site inspection or reasonable suspicion testing, collection of hair specimens from suspected cases was able to carry out under direct supervision without invasion of privacy. Although hair testing was well established as a useful technique in forensic toxicology using chromatography-mass spectrometry methodology^15, 16^, there is an urgent need for a rapid, sensitive, and low-cost method to inspect the hair of suspected drug abusers for on-site inspection. This is particularly challenging using current techniques which typically require complex instruments and a long detection time^13, 17^, and these conditions cannot be met during the law enforcement process.

Lateral flow immunoassay (LFA) is the most popular tool for point-of-care testing (POCT)^18-20^, with the advantages of being rapid, easy to use, and cost-effective. Its applications are widely growing, including for clinical diagnosis^21-23^, food safety^24, 25^, and environmental monitoring^26, 27^. LFA-based drug testing is acceptable for primary public security law enforcement because the process involved is simple and fast. However, the traditional lateral flow assay using colloidal gold labeling has a low sensitivity (100 ng/ml for a urine drug test)^28, 29^, which does not meet the required limit-of-detection for hair drug detection (less than 0.2 ng/mg). Fluorescent nanoparticles, including quantum dots^30-34^, europium chelate-labeled microspheres^23, 35, 36^, upconversion nanoparticles^21, 37, 38^, and aggregation-induced emission nanoparticles^39-41^, have been applied as an alternative approach to improve the detection sensitivity and specificity of various testing and imaging techniques, including LFA-based assays^42-47^. However, these approaches require an excitation light and instrumental reader, which is not reachable at resource-limited setting. Although gold nanoparticle has many advantages including simple synthesis and antibody conjugation procedures^48, 49^, there are still some disadvantages such as non-covalent antibody conjugation, colloidal unsteadiness, intrinsic plasmonics and especially low detection sensitivity. Hence, alternative materials are still sought for naked eye POCT system. Dyed polymer nanobeads have attracted attention as an alternative immunoassay labels^50, 51^. Polymer nanobeads can be prepared from the co-polymerization with dyes, solvent swollen with template nanobeads, and self-assembly of block copolymers and dyes, with variety of surface functional groups^52, 53^. Moreover, polymer nanobeads have excellent biocompatibility and flexibility, which facilitate efficient conjugation of specific proteins and antibodies.

We attempted to improve the sensitivity of lateral flow assay test strip using composite polymer based nanobeads loaded with an increasing quantity of dyes (i.e., deeply dyed) to allow for enhanced label signal and easy target detection using the naked eye. In this study, we encapsulated phthalocyanine dyes in amphiphilic copolymers to prepare a deeply dyed nanobead system with controlled core-shell structure, which was used to develop a highly sensitive nanobeads based lateral flow assay for drug detection. We tested our method for morphine and methamphetamine in hair without complicated sample pretreatment. Our encapsulation approach of loading controlled phthalocyanine dyes in nanobeads, the nanobeads are deep in color, enabling color signal detection with high sensitivity and specificity using the naked eye.

## Experimental

### Materials

Copper (II) phthalocyanine (β-form, dye content 90%) and *N,N’*-Dimethylquinacridone were purchased from Shanghai Macklin Biochemical. Phthalocyanine Green were supplied by Hangzhou Epsilon Chemical Co. Ltd. Poly (styrene-co-maleic anhydride) (PSMA) copolymer, casein, and carbodiimide hydrochloride (EDC) were purchased from Sigma-Aldrich. Sodium dodecyl sulfate (SDS), goat anti-rabbit IgG antibody, and rabbit IgG were purchased from Sangon Biotech. Anti-methamphetamine (MET) antibody, MET-BSA antigen, anti-morphine (MOP) antibody, and MOP-BSA antigen were obtained from Clongene Biotech (Hangzhou, China). The glass fiber paper (SB06) used as sample and conjugated pads, the absorbance pad (CH37 K), and adhesive polyvinyl chloride (PVC) backing plate were obtained from Kinbio Tech (Shanghai, China). Other chemical reagents were obtained from the Sinopharm (Shanghai, China). Positive hair samples were kindly gifted from the third research institute of the ministry of public security (Shanghai, China), and volunteers from our team members provided negative hair samples.

### Preparation of the DDNB

Preparation of DDNB was carried out as follows^30, 34^: 1 mL of chloroform solution containing copper phthalocyanine or similar dyes (5 mg), and matrix polymer PSMA (20 mg) was added into a 5 mL SDS (wt 0.5%) aqueous solution. The polymer/dye doped miniemulsion was prepared by ultrasonication for 60 s (3 s pulse, 3 s pause) at 30% amplitude using a sonicator (Scientz-IID, China) with magnetic stirring at 200 rpm. Afterward, the prepared miniemulsion was solidified by chloroform evaporation overnight in an open container under magnetically stirring. The prepared dyed polymer composite nanobeads were centrifuged at 14,000 rpm for 10 min, washed with deionized water three times, and dispersed in water for storage.

### Characterization of the DDNB

The size and morphology of nanobeads were analyzed with a transmission electron microscope (TEM, JEM-2100F, Japan) at an accelerating voltage of 100 kV and a scanning electronic microscope (SEM, JSM-7401F, JEOL, Japan). A Zetasizer Nano ZS90 instrument (Malvern, England) was used to measure nanobeads’ hydrodynamic diameter in an aqueous suspension. Fourier transforms infrared spectra (FTIR) of the copper phthalocyanine pigment and blue DDNB (in KBr pellet) were recorded on an FTIR spectrometer (Spectrum-100, Perkin Elmer, USA). Adsorption spectra were recorded using an ultraviolet–visible (UV–vis) spectrophotometer (UV-2300, Shimadzu, Japan).

### Antibody conjugation with DDNB

Antibodies were covalently conjugated with DDNB via the carboxyl groups on the surface of DDNB and amino groups in antibodies under EDC activation. Generally, DDNB (100 μl) in 200 μl of phosphate buffer (PB, 20 mM, pH 6.0) and 6 μl of EDC (20 mg/mL) were added into a tube and incubated for 0.5 h to activated carboxyl groups. The activated DDNB suspension was centrifuged at 15,000 rpm for 10 min, and the supernatant with excessive EDC was removed. Then, 200 μL PB buffer was added into the precipitate to disperse by ultrasonication. Anti-MET, anti-MOP, or secondary antibodies (6 μg) were added into 200 μL PB buffer, activated dyed polymer nanobeads were added into antibody solution, and the mixture was incubated at room temperature for 30 min. Excess antibodies were removed by centrifugation at 8,000 rpm for 10 min and decantation of supernatant. 200 μL casein solution (0.5%) was added into the precipitate to disperse by ultrasonication, and unreacted sites on surface of DDNB were blocked at room temperature for 2 h. Finally, a 200 μL preservation solution with preservative (ProClin 300, 0.1%) was added and the DDNB-antibody conjugates stored at 4 °C.

### Fabrication of lateral flow immunoassay strips

The DDNB-based lateral flow immunoassay strip uses a competitive immunoassay format. As shown in **Figure 1**, the test strips composed of a backing plate, sample pad, conjugate pad, NC membrane, and absorption pad. NC membrane were adhered on the backing plate, and MET-BSA or MOP-BSA antigen (0.2 mg/mL) and rabbit IgG (0.5 mg/mL) were dispensed onto the NC membrane in batches, at a jetting rate of 1 μL/cm, to respectively form the test (T) line and control (C) line. The membrane was dried for 24 h at room temperature. The sample pad and absorption pad were used without any treatment. Blue DDNBs were labeled with anti-MET or anti-MOP antibodies, and red DDNBs were labeled with goat anti-rabbit antibodies. DDNB conjugates were dispersed on the conjugate pad, and dried at room temperature for 24 h. Finally, sample pad, conjugate pad, and absorption pad were assembled on the backing plate. The fully assembled backing plates were cut into test strips and stored at desiccative conditions.

**Figure 1.**
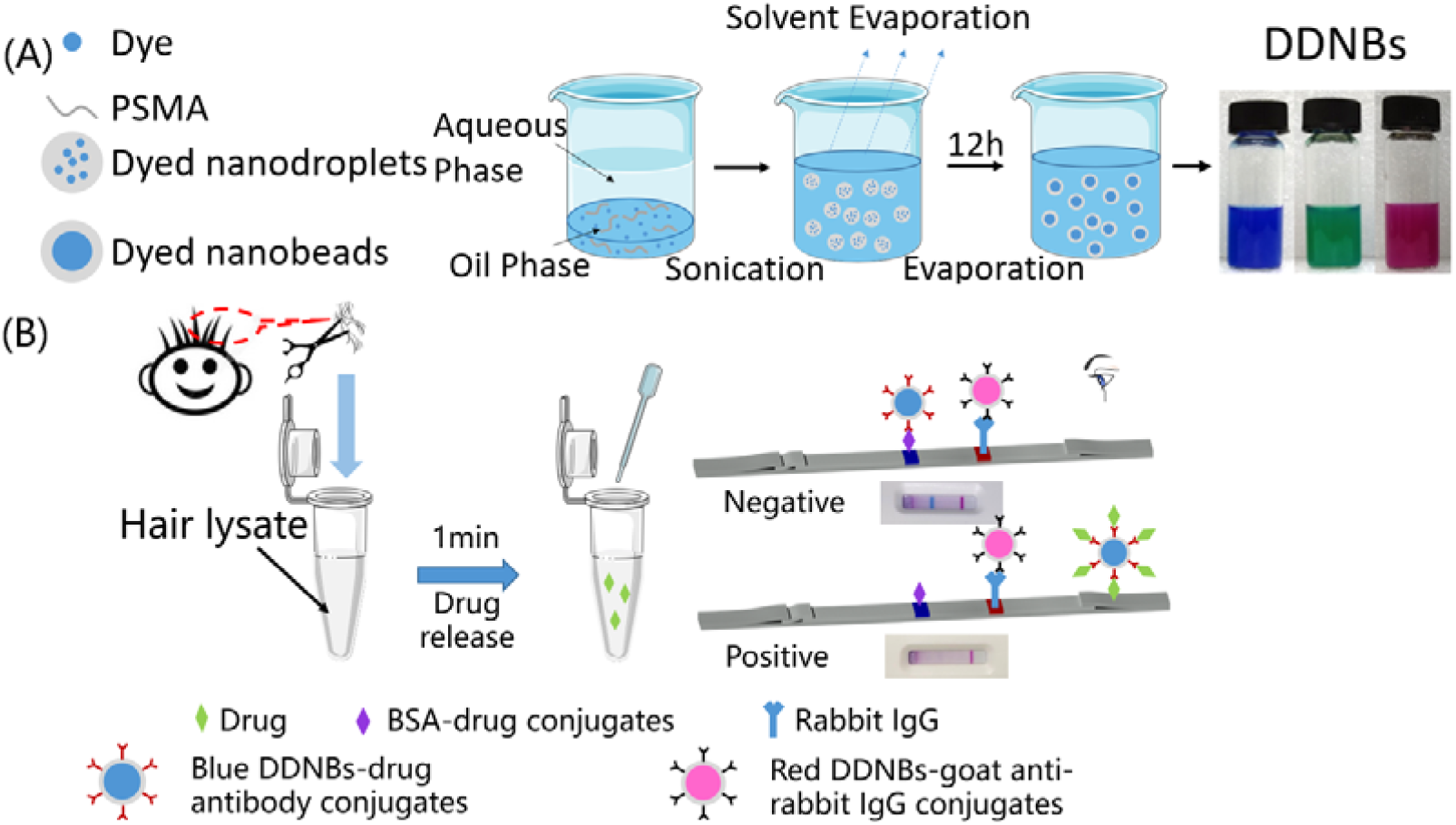
(A) Schematic illustration for the preparation of DDNB and photographs of different color DDNB suspension. (B) The schematic diagram for detecting drugs in hair by a competitive lateral flow immunoassay using dual-color DDNB as labels.

### Testing Procedures for DDNB-LFA

The drug (MOP or MET) calibration solution was prepared by diluting standard drug solution (Sigma-Aldrich) with PBS buffer to a series of different concentration solution, and PBS buffer was used as the blank control. A total of 100 μL of calibration solution was loaded into the sample hole of DDNB-LFA and waiting for desired time (for example, 8 min). For the qualitative detection, the test results were read with naked eye in typical room lighting. For semi-quantitative analysis, color intensity of test line was analyzed using a digital reader (Shenzhen Yuanqin Biotech, China).

Drugs in hair samples were extracted by an enzymatic digestion method. Hair specimens were chopped into 2-5 mm length, milled into powder using a micropipette grinder, and digested in the enzymatic digestion solution (Fuzhou Chuangdian Biotech) for 1 min. Then, 100 μL of the supernatant solution was analyzed by a test strip using aforementioned detection procedures.

## Results and Discussion

### Principle of DDNB-LFA

The deeply dyed nanobeads (DDNB) were prepared via oil/water miniemulsion-solvent evaporation techniques, which were widely used to prepare functional nanobeads by encapsulation of quantum dots^30^, aggregation-induced emission dyes^40^, or magnetic nanoparticles into polymer nanobeads^34^. Amphiphilic copolymer poly(styrene-co-maleic anhydride) (PSMA) was used for the synthesis of dyed composite nanobeads, because it is soluble in chloroform, and carboxyl groups on surface of nanobeads can be obtained by hydrolysis of anhydride groups in PSMA. We choose oil-soluble dyes, including copper phthalocyanine (CuPc), Phthalocyanine Green and *N,N’*-dimethylquinacridone dyes, which have brilliant color, and excellent stability on light, heat, and solvent resistance, have the potential to form highly stable dyed nanobeads. As shown in Figure 1A, the dispersed phase was composed of oil-soluble dyes and PSMA dissolved in chloroform, and emulsifier SDS aqueous solution was used as the continuous phase. Subsequently, an oil/water miniemulsion was obtained by ultrasonication with magnetic stirring. Then, the obtained minidroplets containing both organic dyes and copolymer were solidified by gradual evaporation of chloroform. Finally, the minidroplets solidified into the form of composite nanobeads and entrapped organic dyes inside the polymer nanobeads.

The testing procedures of drugs in hair using the LFA strip were schematically shown in Figure 1B. Firstly, drugs in hair samples were released in buffer via enzymatic digestion. The DDNB-based LFA was based on a competitive immunoassay. Drug-BSA conjugates and rabbit IgG were respectively immobilized on the nitrocellulose membrane as test (T) line and control (C) line. Monoclonal antibody (mAb) against drug was conjugated with blue DDNBs, and goat anti-rabbit IgG was conjugated with red DDNBs. In the sample without the drug, blue DDNB-mAb conjugates were captured by the drug-BSA conjugates immobilized on the T line. While in the sample presence of the drug molecules, DDNB-mAb conjugates were inhibited to bind with drug-BSA conjugates on T line with competing binding of drugs molecules in sample. Thus, optical density of T line was inversely correlated with the drug concentration, and the blue T line were totally disappeared when the drug concentration was high enough. While red DDNBs conjugated with anti-rabbit IgG were captured by rabbit IgG as a control, and the red line was always formed in the control zone with or without a drug in samples.

### Characterization of the prepared deeply dyed nanobeads

As shown in Figure 2A-D, the particle size and core-shell ratio of DDNBs could be controlled by adjusting the ratio of dye (CuPc) to polymer in the oil phase. The average diameters and core-shell ratio of DDNBs measured from TEM images (Figure 2A-D) are listed in Table S1. As shown in Figure 2A-D and Table S1, with constant weight of dyes and polymer in the dispersed phase, as the proportion of the dye increased, the diameter of obtained nanobeads gradually decreased, which was attributed to the relative higher density of CuPc dyes (about 2.8 g/cm^3^) compared with PSMA polymer (1.05 g/cm^3^). With increasing ratio of dye content in the dispersed phase, the core-shell diameter ratio of nanobeads also increases, which means that the shell becomes thinner (as shown in Figures C and D) and more prone to cracking (as shown in Figures S1). However, as the ratio of dyes increased, the color of latex microspheres got darker. To prepare darker latex microspheres under the premise of ensuring the stability of DDNB, we chose the ratio of dye and polymer shown in Figure 2B for antibody conjugation. Figure 2E is the SEM image of the DDNBs prepared at ratio of Figure 2B with a regular spherical and relatively uniform size.

**Figure 2.**
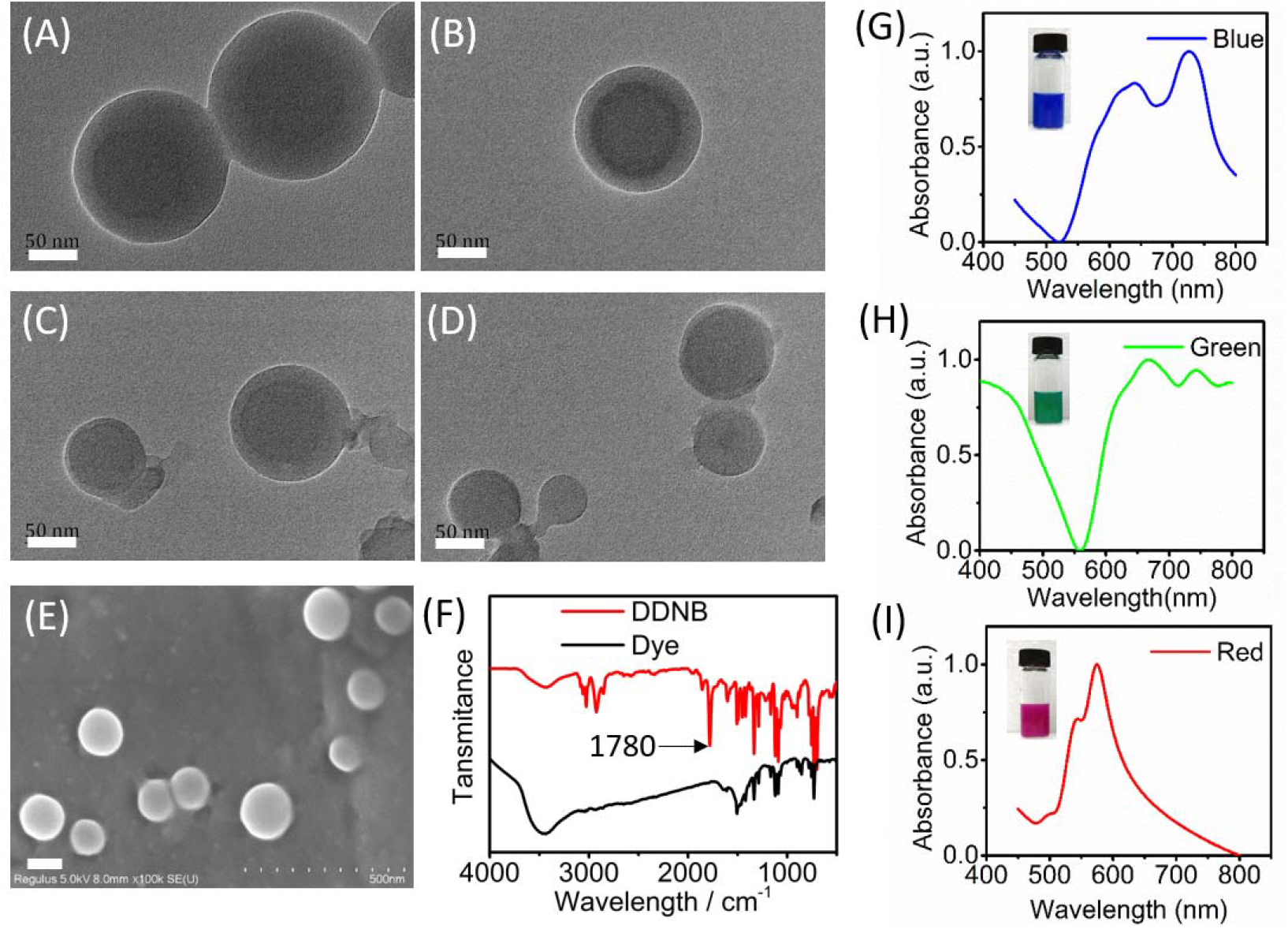
Morphology characterization of DDNB. (A-D) TEM images of DDNB were prepared with different dye/polymer ratios. (E) SEM image of DDNB. The scale bars represent 50 nm in TEM images and 100 nm in SEM images. (F) FTIR spectra of DDNB and copper(II) phthalocyanine dyes. (G-I) UV-vis absorption spectra of DDNB prepared by dyes of different colors. The insets are digital images of the DDNB suspensions tested by UV-vis spectroscopy.

The anhydride groups in PSMA molecules were able to be hydrolyzed in aqueous suspension on the surface of DDNB. As shown in Figure 2F, the FTIR spectra absorption peak around 1780 cm^-1^ in DDNB was assigned to the C=O stretching vibrations in carboxyl groups. Furthermore, red and green color DDNB were also prepared with the same procedures using red dye *N,N’*-dimethylquinacridone and green dye phthalocyanine green. As shown in Figure 2 G-I, the photos of colored DDNB suspensions under normal room lighting had deeply bright colors, and UV-vis spectra of colored DDNB showed different absorbance peaks.

### Stability evaluation of DDNB

The stability of DDNB is essential for preparing LFA probes. To study the stabilities of blue DDNB, the nanobeads suspension was dispersed in different pH buffers (pH 6-9) and high-temperature (60 °C) buffer for a period of time. As shown in Figure 3A, the results showed that the absorbance intensity of the tested nanobeads suspension decreased with an increase of pH value, and the absorbance value decreased by 8.2% from pH 5 to pH 8. However, both acidic and alkaline pH values have an insignificant effect (variation of less than 15%) on the absorbance stability of the latex nanobeads.

**Figure 3.**
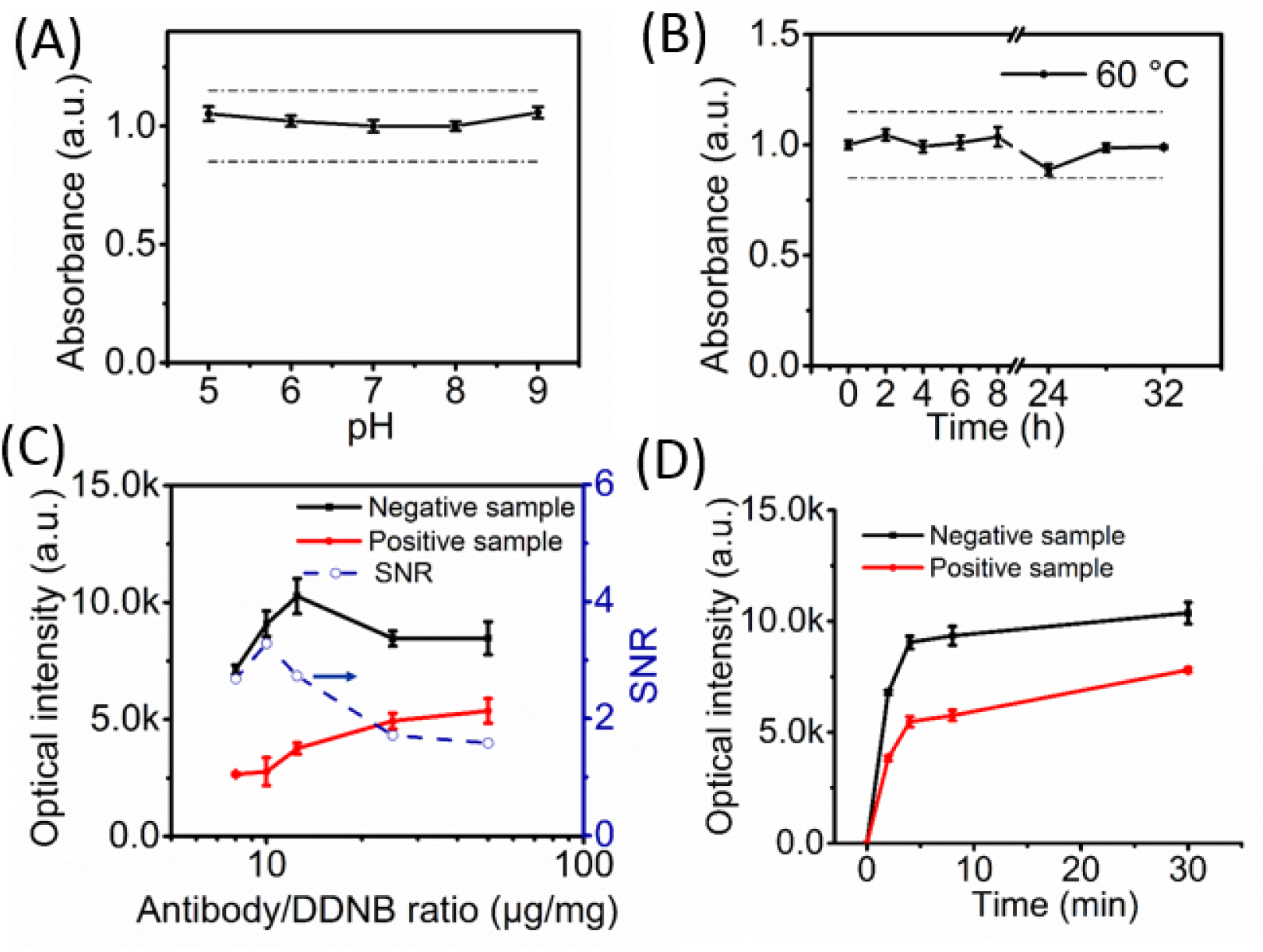
Stability of DDNB suspensions in different pH buffers (A) and at high temperature (60 °C) buffer (B). (Dotted lines in figure A and B represents 15% deviation from the baseline intensity) (C) Effect of antibody/DDNB ratio on the T line optical intensities and signal-to-noise ratio (SNR) of positive and negative samples. (D) Optical intensities of test line developed on the reaction time.

We also tested the stability of the prepared DDNB dispersion at 60 °C for different time points (up to 32 hours), as shown in Figure 3B. The absorbance intensity of DDNB suspensions varied by less than 15%, indicating that our nanobeads were stable at a high temperature of 60 °C.

### Conjugation antibodies with DDNB

To achieve specific recognition of target molecules, DDNB particles were conjugated with anti-drug antibodies. Antibodies with free amino groups were covalently immobilized on the surface of DDNBs, which are rich in carboxyl groups, via carbodiimide (EDC) activation. For small molecules detection, we employed a competitive immunoassay format. DDNB-antibody conjugates were competitively reacted with drug target in a sample and drug-BSA antigen immobilized on the T line. Signal intensity on the T line can be directly affected by the amount of antibody conjugated with DDNB. To achieve a bright blue T line and relatively high sensitivity with minimal background interference for negative samples, antibody/DDNB conjugation ratios were optimized. As shown in Figure 3C, with an increasing antibody/DDNB ratio, the T line intensity gradually increased for positive samples, while for negative samples, the peak T line intensity occured at 15 μg/mg of antibody/DDNB ratio. Continuously increasing antibody/DDNB ratio slightly decreased the T line intensity, which may be attributed to excessive antibodies located on DDNB surface saturating the binding sites of drug antigen, and reducing the number of DDNB binding on T line. However, the signal/noise ratio (SNR) calculated from the T intensity of positive and negative samples peaked at 15 μg/mg of antibody/DDNB ratio.

Using the optimized antibody/DDNB ratio, we studied the influence of reaction time on the T and C line intensity of the DDNB-LFA system. As shown in Figure 3D, although the T and C line intensity nearly reached the plateau at 5 min, we performed further detection procedures until 8 min to reduce variation of T line intensity with reaction time.

### Performance evaluation of DDNB-LFA

The sensitivity of DDNB-LFA was compared with gold nanoparticle base LFA (GNP-LFA) by testing different concentrations of MET calibration solution. For the DDNB-LFA system, we labeled MET-mAb with the blue colored DDNB and goat anti-rabbit IgG with red colored DDNB. Thus, the T line coated with MET-BSA antigen displayed a blue line for negative samples, and the T line displayed a weak blue line or disappeared for positive samples. The C line consistently displayed a red line for all samples. For the qualitative detection of MET, the minimum MET concentration corresponding to the strip with an absent T line was defined as the cut-off concentration for the MET test. As shown in Figure 4A, the intensity of the blue T line gradually decreased with the MET concentration reached 1.0 ng/mL, and the T line entirely disappeared at 8.0 ng/mL. While for the GNP-LFA system, the T line entirely disappeared at 25 ng/mL, which was about 3.1 times less sensitive than the DDNB-LFA system.

**Figure 4.**
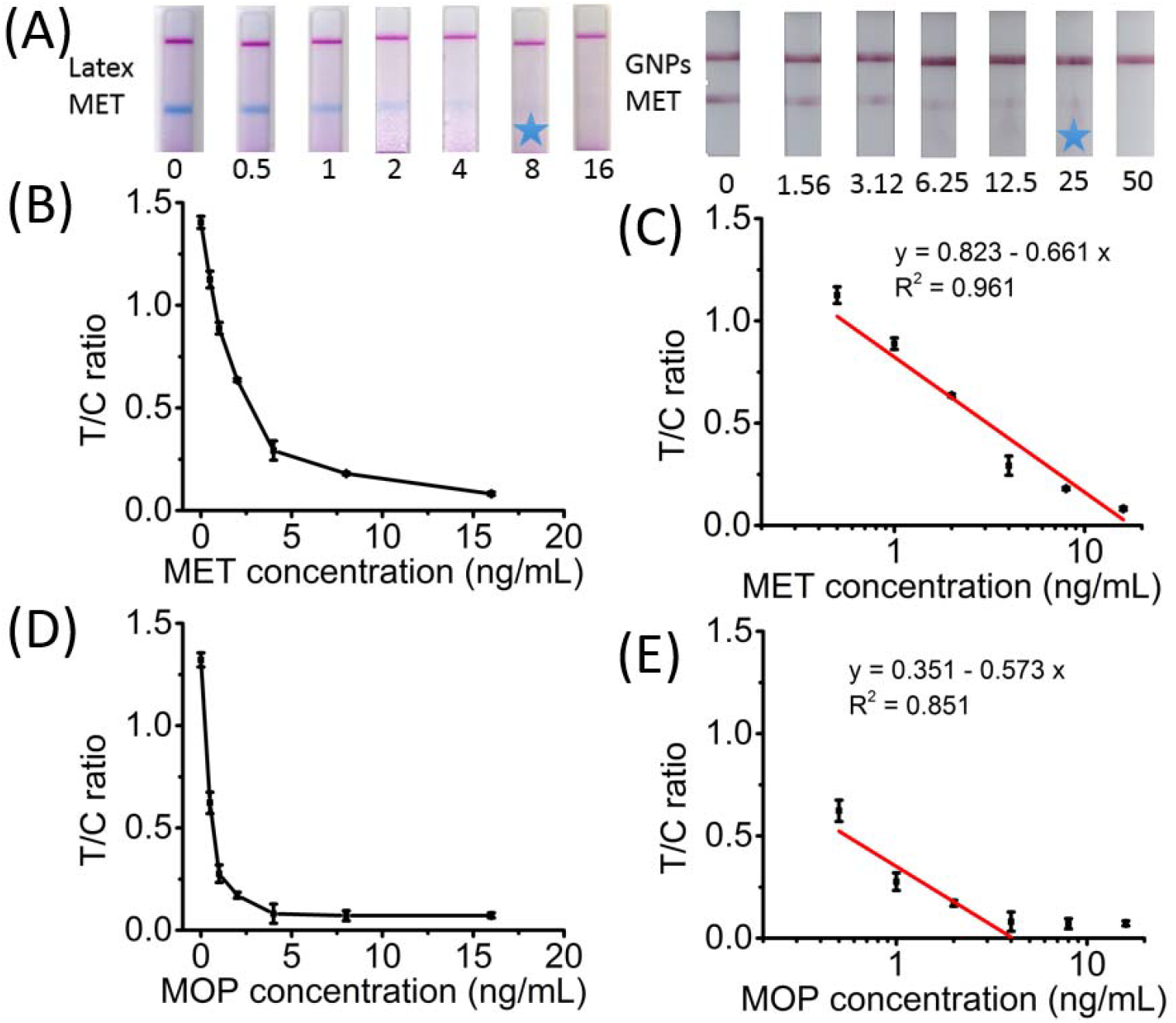
(A) Comparison of the detection sensitivity of DDNB-LFA and GNP-LFA system for the qualitative detection of MET with naked eye. The cut-off concentration of immunoassay systems was marked with a blue star. A standard calibration curve (B) and linear regression (C) after logit transformation to detect MET calibration solution using DDNB−LFA system. A standard calibration curve (D) and linear regression (E) after logit transformation to detect MOP calibration solution using DDNB−LFA system.

Semi-qualitative test results of DDNB-LFA assay were obtained from a digital strip reader and was recorded after running for 8 min. The resulting calibration plot of the drug assay, as shown in Figure 4B and 4D, showed that the T/C ratios were inversely related to the drug concentration. As shown in Figures 4C and 4E, experimental data indicated a good linear relationship between T/C ratio and log [concentration of drug], ranging from 0.5 to 16 ng/mL for MET calibration solution and from 0.5 to 4 ng/mL for MOP calibration solution.

The specificity of the developed DDNB-LFA system for MET detection was investigated using a series of interference drugs, including morphine, cefprozil, levofloxacin, aspirin, and ibuprofen. Artificial MET positive (10 ng/mL) and negative MET samples spiked with interfering drugs (100 ng/mL) were tested, as shown in Figure 5A. The results showed that the presence of the tested non-target drugs (morphine, cefprozil, levofloxacin, aspirin, and ibuprofen) have an insignificant effect (*p* > 0.05) on the detection of MET using the developed DDNB-LFA system even at high concentration (100 ng/mL) of interfering drugs.

**Figure 5.**
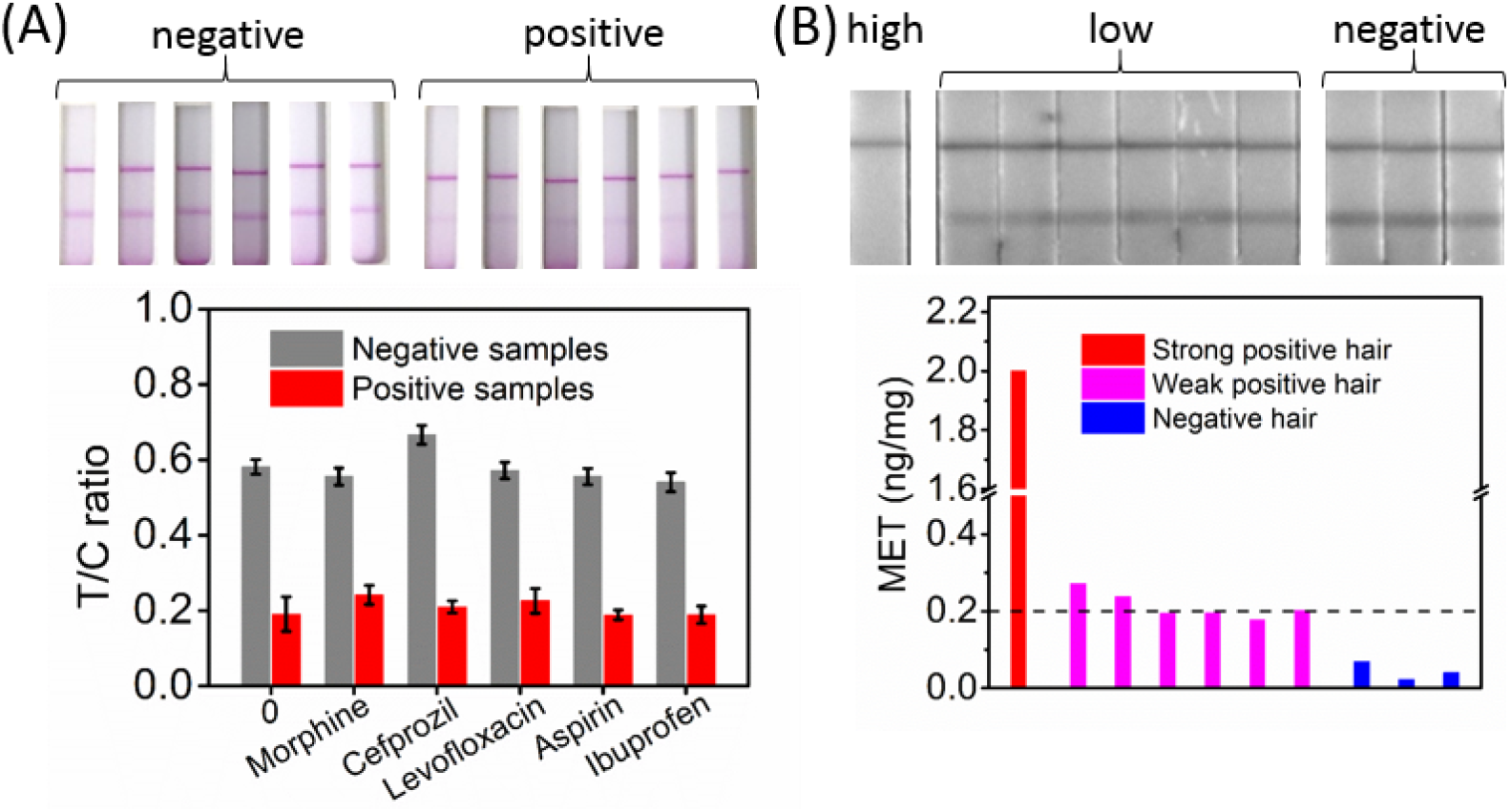
(A) Specificity of DDNB-LFA for MET detection. The concentration of interfering drugs, including morphine, cefprozil, levofloxacin, aspirin, ibuprofen, was 100 ng/mL. (B) Detection results of MET concentration in human hair samples through DDNB-LFA system.

### Hair sample testing

To evaluate the performance of the developed DDNB-LFA method for rapid drug testing in hair samples. We tested MET in hair samples with different doses of drug pre-determined by liquid chromatography-mass spectrometry (LC-MS), including three negative samples, six weak positive samples (about 0.2 ng/mg), and one strong positive (> 2 ng/mg) sample. The hair sample (5 mg) were chopped into 3-5 mm lengths, milled into ultrafine powder, and extracted with the enzymatic hair lysate buffer (500 μL) for 1 min. Enzymes can be used to promote the extraction of drugs from hair samples. After 1-3 min incubation, 100 μL of lysate buffer extracted with hair sample was loaded into the DDNB-LFA system and run for 8 min. As shown in Figure 5B, the results indicated that the developed DDNB-LFA system was practical for accurate hair sample testing. The test results of the developed DDNB-LFA system were consistent with the results measured by the LC-MS method.

## Conclusions

In summary, we have developed a deeply dyed nanobeads system for sensitive naked eye lateral flow immunoassay. The prepared dyed nanobeads displayed controlled core-shell structure, stable absorbance signals and easy for antibody conjugation. These deeply dyed nanobeads were more sensitive than traditional gold nanoparticles based lateral flow immunoassay. Moreover, these colorful dyed nanobeads are potential for multiplexed immunoassay strips for simultaneous multiple analytes detection.

## Data Availability

All data produced in the present work are contained in the manuscript

## Acknowledgments

This work was supported by the Project of Shanghai Science and Technology Committee (STCSM) (20S31902800, 19441904200), the International Science & Technology Cooperation Program of China (2014DFA33010), and partially supported by National Key Research and Development Program of China No. 2017YFC0803606.

## Appendix A. Supplementary data

Supplementary data related to this article can be found online.

